# ANTI-CITRULLINATED ALBUMIN ANTIBODIES AS BIOMARKER FOR THE DIAGNOSIS OF RHEUMATOID ARTHRITIS

**DOI:** 10.1101/2023.08.04.23292887

**Authors:** Vishnupriya R. Paturi, Ramakrishna R. Uppuluri, Lina Gao, Charles T. Roberts, Srinivasa R. Nagalla

## Abstract

**Objective:** Citrullinated human serum albumin has been previously reported in serum and synovial fluid from individuals with rheumatoid arthritis (RA), and antibodies to citrullinated human serum albumin (ACA) have been identified in RA serum. We developed a point-of-care (POC) test for ACA and evaluated its sensitivity and specificity in healthy subjects and subjects with clinically diagnosed RA and other rheumatic conditions and autoimmune disease.

**Methods:** The POC test is a lateral-flow immunoassay using antihuman IgA/G/M and anti-human serum albumin antibodies for detection of citrullinated serum albumin-ACA complexes. This test was used to determine serum or plasma ACA levels in a South Asian study population comprised of healthy controls (n=484) and subjects with clinically diagnosed RA (n=354) or other rheumatic (n=103) and autoimmune diseases (n=60), and compared to the levels of rheumatoid factor (RF) and anticitrullinated cyclic peptide antibodies (ACPA).

**Results:** Sensitivity of ACA for RA was 0.520 and specificity was 0.994. ACA prevalence in other rheumatoid disease was similar to that of ACPA and less than that of RF. ACA was detected in 12% of RA samples that were negative for ACPA. The combined sensitivity of ACA+ACPA was 0.664 and the combined specificity was 0.845.

**Conclusion:** The ACA POC test exhibits robust sensitivity and specificity for RA diagnosis in serum or plasma and, in conjunction with ACPA, can enable rapid and efficient differential diagnosis of RA with increased sensitivity and comparable specificity.

## INTRODUCTION

Rheumatoid arthritis (RA) is an autoimmune disease with unclear etiology, and is characterized by joint inflammation and underlying bone loss [1]. RA preferentially affects women [2], and its global prevalence is 1% [1,3]. RA diagnosis is typically accomplished through a combination of clinical assessments that are complemented by determination of the presence of specific biomarkers [4]. RA biomarkers are also potentially important in the identification of at-risk individuals prior to the development of clinically apparent disease [5,6] in order to institute prophylactic therapies, including new approaches targeting specific inflammatory mediators [7]. The principal RA biomarkers are rheumatoid factor (RF), which is comprised of antibodies against the Fc domain of IgG, and anti-citrullinated peptide antibodies (ACPAs) [8] that recognize proteins containing arginine residues that have been deimidated, or citrullinated [9] by members of the peptidyl arginine deimidase (PAD) family [10,11]. This modification is thought to render the citrullinated proteins more immunogenic, resulting in more frequent autoimmune responses [12]. Although RF is detectable in the majority of RA patients, it is also seen in other connective tissue diseases such as systemic lupus erythematosus (SLE) and Sjogren’s Syndrome (SS) [13]. Although there is recent evidence that the epitopes recognized by RF and ACPA antibodies may be shared in some cases [14], ACPAs appear to exhibit more specificity for RA than RF (i.e., up to 80% in serum of RA patients) [15], although ACPAs, like RF, are also detectable in non-RA conditions such as SLE and SS [16]. ACPA is the primary biomarker currently employed in RA diagnosis.

A number of studies have characterized a spectrum of citrullinated proteins (“citrullinome”) in synovial fluid [17-20] and serum [20] of RA patients. A subset of antibodies against these proteins is what is detected by the standard ACPA test. Citrullinated human serum albumin (HSA) in particular was identified in RA synovial fluid [18-20] and serum [20], and Hefton et al. subsequently reported the presence of anti-citrullinated HSA antibodies (ACA) in RA serum [21]. In that study of 79 RA patients, 38% had ACA levels >2 SD’s above healthy controls, and ACA positivity trended higher in females and in patients with joint erosions. In the present study, we extend these observations using a newly developed point-of-care (POC) immunoassay for ACA in serum or plasma and compared its performance to standard ACPA or RF assays in a large East Asian study population.

## METHODS

During the development of a prototype lateral-flow immunoassay for SARS-CoV-2 antibodies [22] that incorporated HSA detection in the test line, we detected a positive reaction with RA samples. Subsequent investigation determined that the test line positivity reflected the presence of citrullinated HSA-ACA complexes in the RA samples, similar to the earlier finding of Hefton et al. [17]. We subsequently designed a lateral-flow immunochromatographic test (described below) to directly assess the levels of ACA in serum plasma.

### Anti-citrullinated HSA antibody lateral-flow POC test

Serum or plasma samples were analyzed using an ACA POC test system (Diabetomics, Inc, Hillsboro, OR). The ACA test system consists of an immunochromatography test strip in a cassette and a portable reader for quantification. Test strips are configured with antibodies against human IgG, IgM, and IgA and human serum albumin. Colloidal gold was used as detection reagent. Streptavidin coupled to colloidal gold and biotin on the control line. 150 μl of 1:80 diluted serum or plasma is added to the test strip and inserted into a proprietary reader that utilizes image analyses for quantification and a QRcode recognition module for test and lot-specific information for calibration and quantification. The ACA concentration is displayed at the end of 15 minutes. Results are reported as arbitrary Units/mL, with a value >20 considered positive (>2 SDs above the range seen in healthy controls). The dynamic range of the ACA assay is 15-4,000 U/mL, and intra- and inter-assay coefficients of variation are 6.2 and 9.8%, respectively.

### ACA in subjects with rheumatic and other autoimmune diseases

In a prospective study, serum or plasma samples were collected from 354 subjects with clinically diagnosed RA, 103 with other rheumatic disease (non-RA; 25 SLE, 22 spondyloarthritis, 11 gout, 8 psoriatic arthritis, 7 osteoarthritis, 2 ankylosing spondylitis, 1 polyarthritis, and 27 non-RA rheumatic disease samples of undefined category), and 60 with other autoimmune disease (20 Graves’ disease, 10 myasthenia gravis, 20 GADA+ type-1 diabetes, and 10 chronic kidney disease). Samples were tested with the ACA POC device described above as well as with commercial assays for RF and ACPA (RF and anti-CCP IgG assays from Siemens Healthcare Diagnostics; positive values are >12 IU/ml and >5 U/ml, respectively). In a separate cross-sectional study, 484 banked serum (334) or plasma (150) samples from healthy controls (HC) were tested for ACA. Institutional IRB approval was obtained, the study was registered with regulatory authorities (CTRI/2020/10/028596), and informed consent was obtained from all study participants.

### Statistical analyses

Pair-wise comparisons of ACA levels in the different sample groups employed the Wilcoxon rank sum test followed by Bonferroni adjustment. ACA levels in female vs male RA and non-RA samples were analyzed using the Wilcoxon rank sum test. Age differences in ACA levels were analyzed using one-way ANOVA.

## RESULTS

**Figure 1A** shows ACA levels in HC, RA, and non-RA subjects. Intergroup differences were significant in each pairwise comparison (p<0.0001). At a positive value cut-off of >20 U/mL, 3 of 484 HC subjects (0.6%), 184 of 354 (52%) of RA subjects, and 12 (1 gout, 1 ankylosing spondylitis, 1 osteoarthritis, 1 spondyloarthritis, 6 SLE, and 2 non-RA uncategorized rheumatic disease samples) of 103 (12%) non-RA subjects were positive for ACA. From these data, the calculated sensitivity of ACA for RA was 0.520 and the specificity was 0.996. **Figure 1B** shows ACA levels in the 184 ACA+ (>20 U/mL) RA and 12 ACA+ non-RA subjects. The difference in ACA levels in the ACA+ RA and non-RA groups was also significant (p=0.019). In a separate analysis, all 60 samples from patients with Graves’ disease, myasthenia gravis, type-1 diabetes, and chronic kidney disease were negative for ACA (data not shown).

**Figure 1.**
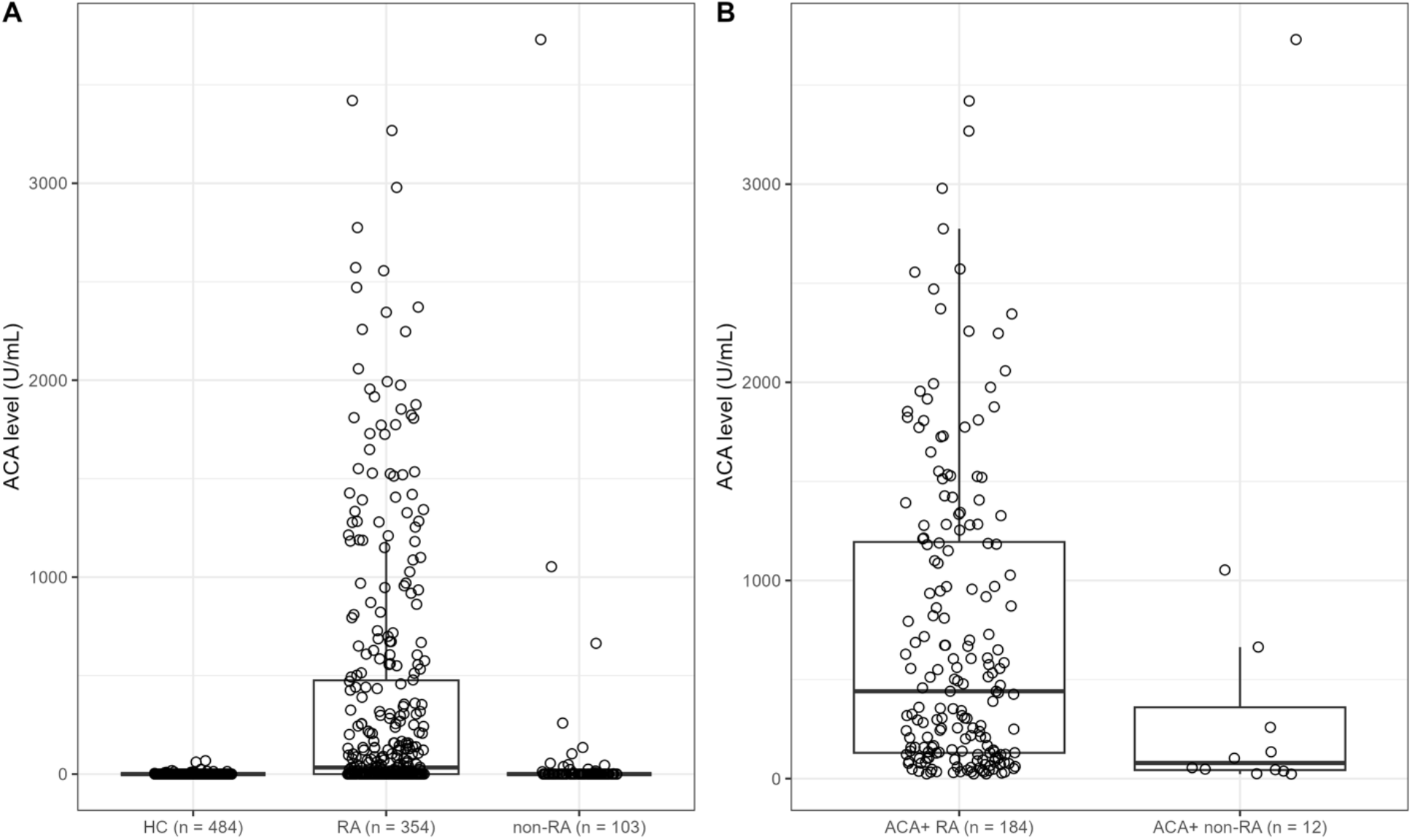
ACA levels in healthy controls (HC) and in RA and non-RA samples. A. ACA levels on all subjects in each group. Kruskal-Wallis test p-values were <0.0001 in all 3 pairwise comparisons (Wilcoxon rank sum test followed by Bonferroni adjustment). B. ACA levels in ACA+ (> 20 U/ml) RA and non-RA subjects. In this comparison, the Wilcoxon rank sum test p-value was 0.019.

We also determined the prevalence of ACPA and RF in the non-RA group. Of the 103 non-RA samples, 8 (8%) were positive for ACPA (3 SLE, 2 gout, 1 polyarthritis, and 2 uncategorized rheumatic disease), and 24 (23%) were positive for RF (3 gout, 1 polyarthritis, 11 SLE, 4 spondyloarthritis, and 7 uncategorized rheumatic disease). Three of these SLE samples and one of the gout samples were positive for all three biomarkers. Thus, RF exhibited the most cross-reactivity with non-RA rheumatic disease samples in this sample set. The sensitivity and specificity of ACA, ACPA, and RF in the RA and non-RA sample sets are summarized in **Table 1**. ACPA and RF values were not available for the HC and other autoimmune disease groups, so ACA specificity in the non-RA group is shown for comparison to the specificity determinations for ACPA and RF using the non-RA group data.

**Table 1.**
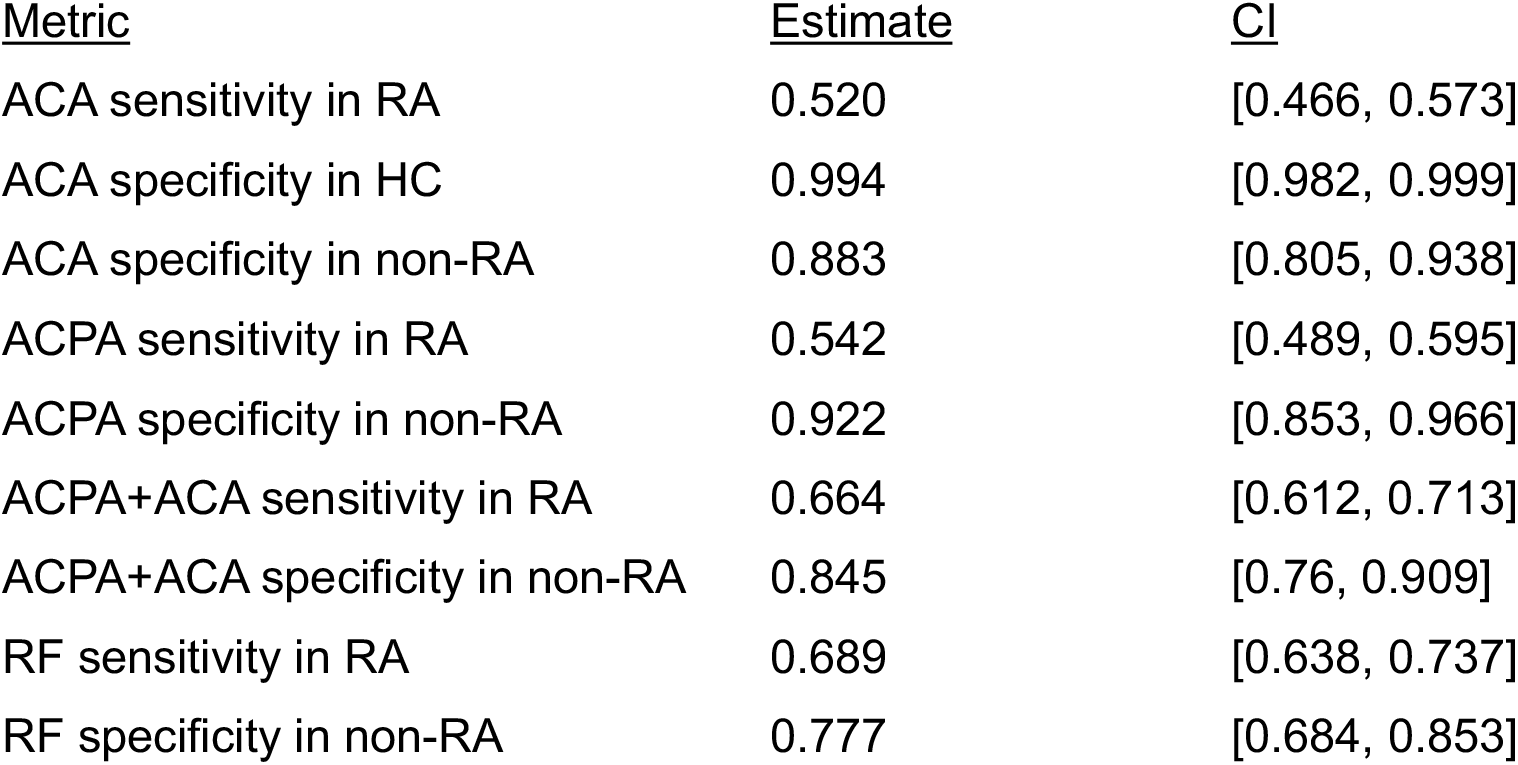
Sensitivity and specificity of ACA, ACPA, and RF. Exact binomial confidence limits (95% CI) are shown.

Since ACPA is the primary biomarker currently employed for RA diagnosis, we examined further the relationship between ACA and ACPA status in the RA sample set (n=354). As illustrated in **Figure 2**, 235 (66%) were positive for either ACPA or ACA. 141 (40%) were positive for both, while 43 (12%) were positive for ACA alone and another 51/354 (14%) were positive for or ACPA alone. Thus, the addition of ACA to ACPA increased the percentage of biomarker-positive samples from 54% to 66% (192 to 235).

**Figure 2.**
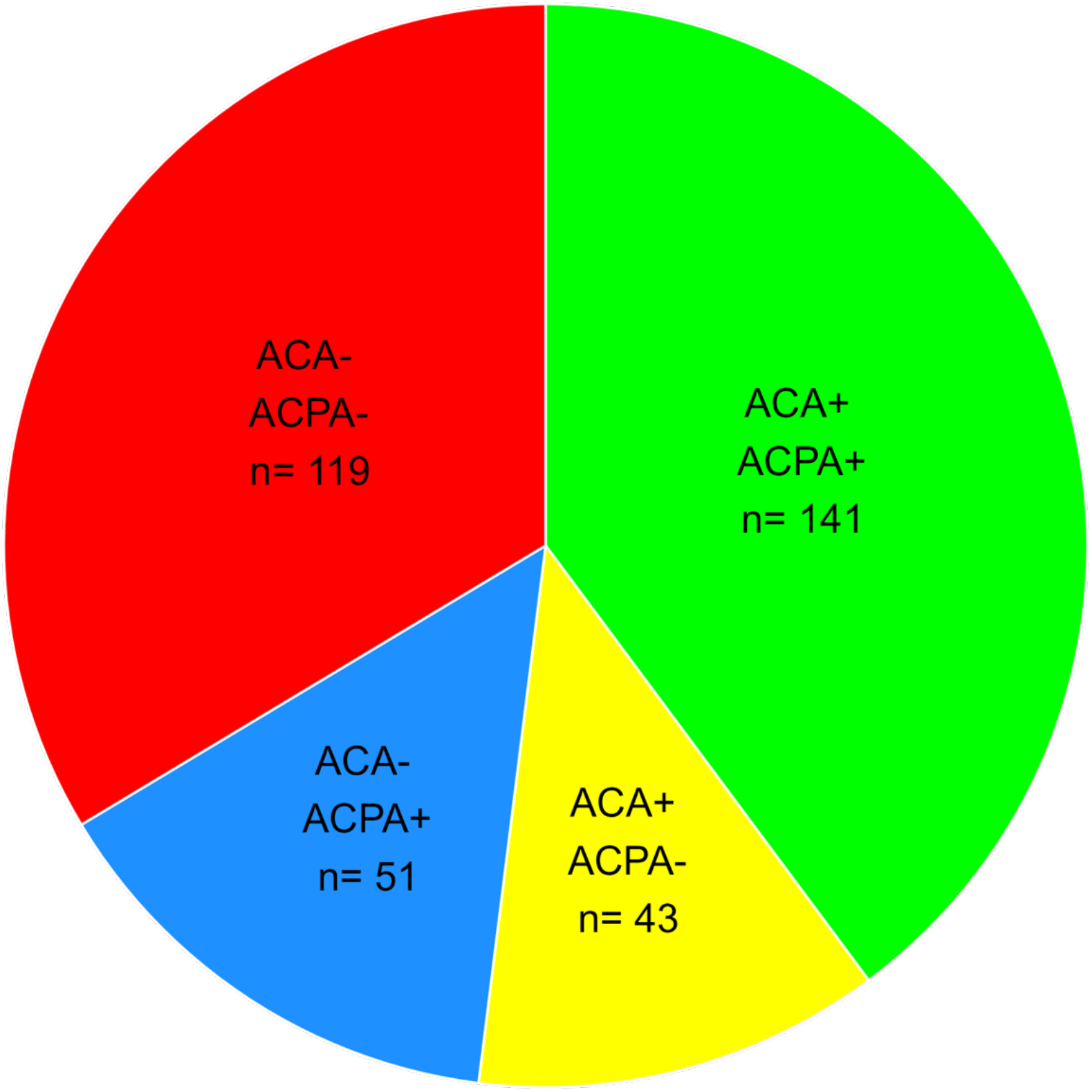
ACA specificity and relationship to ACPA.

We also determined if there was any sex or age difference in ACA levels in the RA and non-RA groups. **Figure 3A** shows the sex distribution of ACA levels in the RA and non-RA groups. ACA levels were significantly higher in females (Wilcoxon rank sum test p-value = 0.0411) in the overall RA group, but not in the non-RA group (Wilcoxon rank sum test p-value = 0.205). This is similar to the trend of higher ACA levels in female RA subjects in the US cohort reported previously [21]. Sensitivity of ACA in females was 0.540 [CI, 0.483, 0.597] and specificity was 0.877 [CI, 0.772, 0.945], similar to the ACA sensitivity and specificity in the total cohort (i.e., 0.520 and 0.883 (ACA specificity in non-RA group), respectively. ACA sensitivity in males was lower that seen in the overall group (0.372 vs 0.520), while specificity was similar (0.892 vs 0.887). However, as shown in **Figure 3B**, when the analysis was restricted to ACA+ samples, no sex difference was apparent (p=0.66 in RA group and p=0.497 in non-RA group).

**Figure 3.**
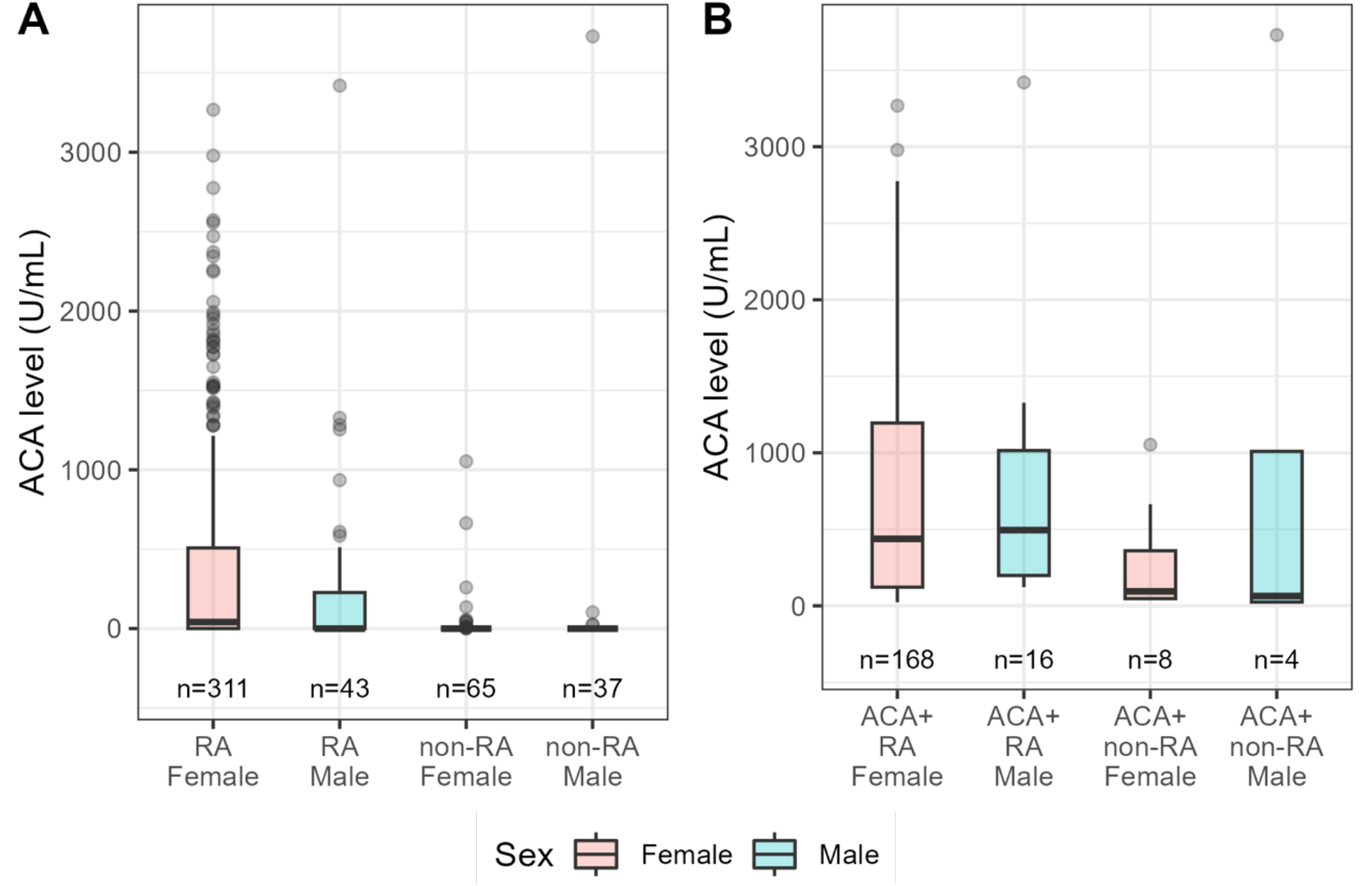
ACA levels in female and male subjects in the overall RA and non-RA groups (A.) and ACA+ RA and non-RA groups (B).

As shown in **Figure 4**, there were no significant differences in age between the various biomarker combinations (one-way ANOVA p-value= 0.526).

**Figure 4.**
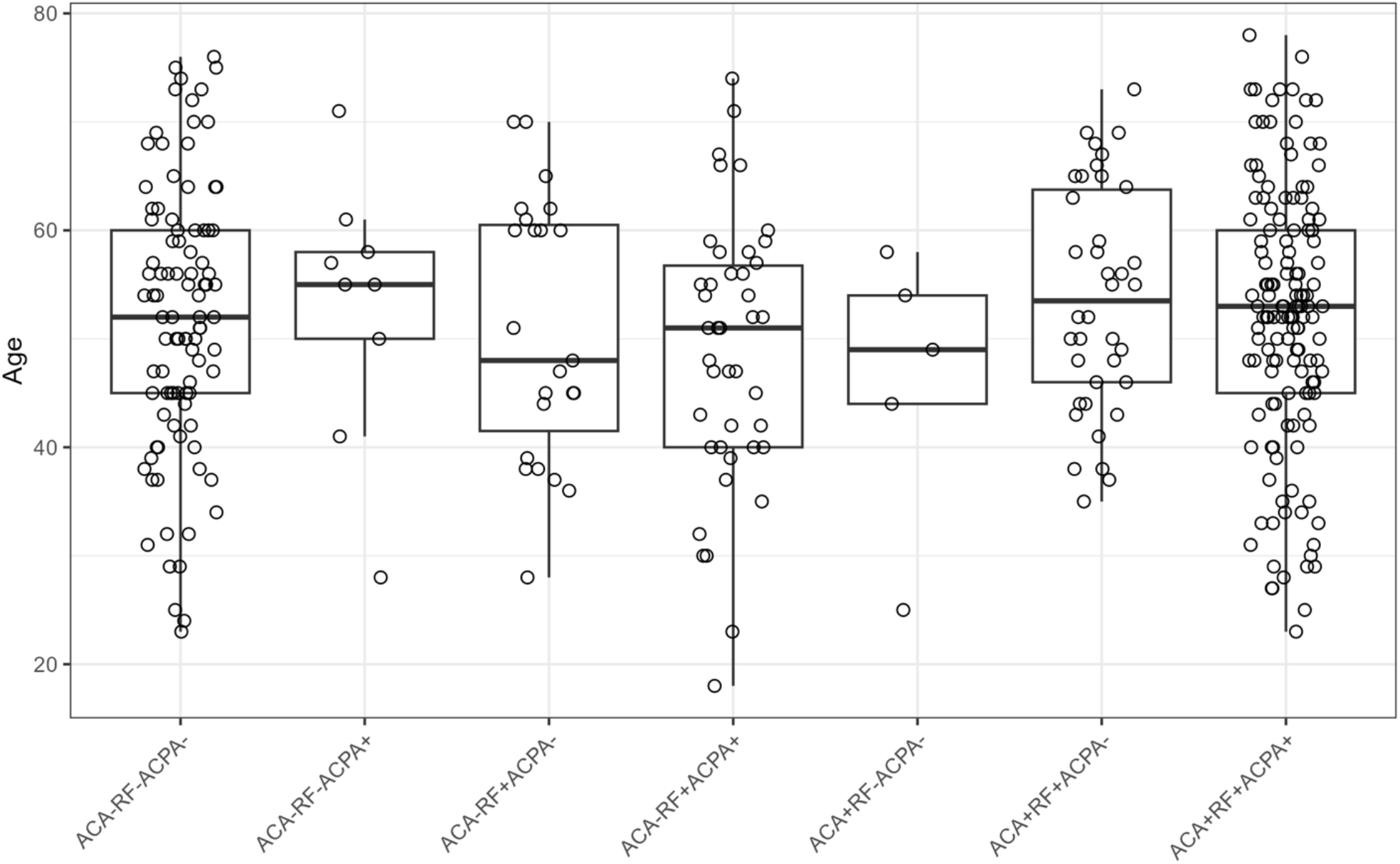
Age distribution in all biomarker combinations in the RA group.

## DISCUSSION

RA is diagnosed and distinguished from other rheumatoid diseases through a combination of clinical assessments and the presence of elevated blood levels of biomarkers, principally ACPA. In the course of development of a rapid POC test for SARS-CoV-2 antibodies, we observed a positive reaction with RA serum samples that was determined to result from the presence of ACA, as previously reported by Hefton et al. [21]. The presence of citrullinated HSA in the circulation and the subsequent generation of ACA may be the result of leakage of citrullinated HSA from synovial fluid or direct citrullination of circulating HSA by PAD activity. PADs are a critical component of neutrophil extracellular trap (NET) formation (NETosis), which is a significant component of RA pathology [22]. Indeed, it was previously shown that the PAD4 isoform implicated in HSA citrullination [21] is released during Neotses’ from neutrophils in synovial fluid [23] and presumably in the circulation as well, providing a plausible mechanism for generating citrullinated HSA in the circulation and subsequent generation of ACA.

These findings prompted us to develop a lateral-flow immunochromatographic POC test to detect ACA. The resulting ACA POC exhibited excellent sensitivity (0.520). and specificity (0.996) in a large East Asian sample set comprised of healthy controls, clinically diagnosed RA cases, and other rheumatoid and autoimmune diseases. ACA levels detected with this test were present in a small percentage (14%) of non-RA rheumatic disease samples, vs 8 and 23% for ACPA and RF, respectively. Importantly, a significant number of RA samples were positive for ACA but not for ACPA. In this sample set, the addition of ACA to ACPA increased the detection of RA from 54 to 66%, thereby increasing overall sensitivity. The addition of an ACPA test line to the current ACA POC would provide a convenient and comprehensive assessment of RA status that would be particularly useful in low-resource environments. Wang et al. [24] have recently described the use of the POC ACA test described here in both serum and plasma samples from a US cohort, and also found that ACA was highly specific for RA.

## Data Availability

All data produced in the present work are contained in the manuscript.

